# Detecting Emerging COVID-19 Community Outbreaks at High Spatiotemporal Resolution — New York City, June–July 2020

**DOI:** 10.1101/2020.07.18.20156901

**Authors:** Sharon K. Greene, Eric R. Peterson, Dominique Balan, Lucretia Jones, Gretchen M. Culp, Annie D. Fine, Martin Kulldorff

## Abstract

New York City’s Health Department developed a SARS-CoV-2 percent test positivity cluster detection system using census tract resolution and the SaTScan prospective space-time scan statistic. One cluster led to identifying a gathering with inadequate social distancing where viral transmission likely occurred, and another cluster prompted targeted community testing and outreach.

---

Spatiotemporal analysis of high resolution COVID-19 data can support local health officials to monitor disease spread and target interventions (*1,2*). Publicly available data have been used to detect COVID-19 space-time clusters at county and daily resolution across the US (*3,4*) and purely spatial clusters at ZIP code resolution in New York City (NYC) (*5*).

For routine public health surveillance, the NYC Department of Health and Mental Hygiene (DOHMH) uses the case-only space-time permutation scan statistic (*6*) in SaTScan^1^ to detect new outbreaks of reportable diseases (*7*) (e.g., Legionnaires’ disease (*8*) and salmonellosis (*9*)). Given wide variability in SARS-CoV-2 testing across space and time, case-only analyses could be poorly suited for COVID-19 monitoring, as true differences in disease rates would be indistinguishable from changes in testing rates across space and time. Moreover, we sought to detect newly emerging or re-emerging hotspots during an ongoing epidemic, which is more challenging than detecting a newly emerging outbreak in the context of minimal or stable disease incidence.

A new approach was needed to detect areas where COVID-19 diagnoses were increasing or not decreasing as quickly relative to other parts of the city. We launched the system on June 11, 2020 to detect community-based clusters of increased SARS-CoV-2 test positivity in near-real time at census tract resolution in NYC, accounting for testing variability.

## The Study

Clinical and commercial laboratories are required to report all results (including positive, negative, and indeterminate results) for SARS-CoV-2 PCR tests for New York State residents to the New York State Electronic Clinical Laboratory Reporting System (ECLRS) (*10*). For NYC residents, ECLRS transmits reports to DOHMH. Laboratory reports include specimen collection date and patient demographics, including residential address. Patient symptoms and illness onset date, if any, are obtained through patient interviews, although not all patients are interviewed.

To detect emerging clusters, the space-time scan statistic uses a cylinder where the circular base covers a geographical area and the height corresponds to time (*11*). This cylinder is moved, or “scanned,” over both space and time to cover different areas and time periods. At each position, the number of cases inside the cylinder is compared with the expected count under the null hypothesis of no clusters using a likelihood function, and the position with the maximum likelihood is the primary candidate for a cluster. The statistical significance of this cluster is then evaluated, adjusting for the multiple testing inherent in the many cylinder positions evaluated.

To quickly detect emerging hotspots, prospective analyses are conducted daily (*12*). To adjust for the multiple testing stemming from daily analyses, recurrence intervals are used instead of p-values (*13*). A recurrence interval of D days means that under the null hypothesis, if we conduct the analysis repeatedly over D days, then the expected number of clusters of the same or larger magnitude is one.

The space-time scan statistic can be utilized with different probability models. We used the Poisson model (*11*), where the number of cases is distributed according to the Poisson probability model, with an expected count proportional to the number of persons tested. Analyses were adjusted non-parametrically for purely geographical variations that were consistent over time, as the goal was to detect newly emerging hotspots. Fitting a log-linear function, we also adjusted for citywide temporal trends in percent positivity, as the goal was to detect local hotspots rather than general citywide trends. To prioritize quickly emerging clusters to identify epidemiologic linkages, we used a short maximum temporal window of 7 days. As we wished to also detect sustained clusters to inform place-based resource allocation, we additionally ran analyses using a maximum temporal window of 21 days.

We developed SAS code (SAS Institute, Inc., Cary, NC, USA) that generated input and parameter files (Table 1, Technical Appendix Table 1), invoked SaTScan in batch mode, read analysis results back into SAS for further processing, and output files to secured folders, including a patient linelist, visualizations, and investigator notification email. Similar SAS code referencing markedly different input parameters is freely available.^2^ Parameters were adjusted during the study period to improve signal prioritization, including increasing the minimum number of cases in clusters from 2 to 5 cases.

**Table 1.** Input file specifications for SARS-CoV-2 percent positivity analyses in New York City, using the prospective Poisson-based space-time scan statistic.

| Feature | Selection | Notes |
| --- | --- | --- |
| Geographic aggregation | Census tract (defined using US Census 2010 boundaries) of residential address at time of report | With less aggregated data, the more precisely areas with elevated rates can be identified. New York City has 2165 census tracts. If geocoding is infeasible, then ZIP Code could be used, but with a loss of spatial precision. |
| Case file | Unique persons reported with a positive result for a molecular amplification detection (PCR) test for SARS-CoV-2 RNA in a clinical specimen. Retain specimen collection date of first positive test. | Confirmed COVID-19 cases* |
| Population file | Unique persons reported with a molecular amplification detection (PCR) test for SARS-CoV-2 RNA in a clinical specimen. For persons who ever tested positive, retain specimen collection date of first positive test. Otherwise, retain most recent specimen collection date. For a given census tract and date, if no specimens were collected, then include in file as having zero population. | Necessary to control for spatial and temporal variability to testing access. We do not use a Census-based population denominator because with a numerator of testing positive conditional on having been tested and a total population denominator unconditional on testing, results would have been difficult to interpret. |
| Omissions from input files | Residents of long-term care facilities, correctional facilities, facilities housing people with developmental disabilities, or homeless shelters; persons whose home address matches a provider or facility; persons diagnosed in the 14 days prior to a more recent case residing in the same building identification number from geocoding; persons with COVID-19 illness onset (where available from patient interview) prior to first date of study period. | To focus on detecting recent community-based transmission, exclude: residents of congregate settings, because building-level clusters are detected using other methods;‡ persons whose listed home address is not a residence; >1 case per building; patients diagnosed long after illness onset. |
| Date of interest for analysis | Specimen collection date | Defining reportable disease clusters according to when patients became ill is preferred. Specimen collection date is the earliest date |
|  |  | available for the study population of persons tested. |
| Study period | 21 days† for analysis to support prioritization of case investigations; since June 1, 2020 for analysis to support place-based resource allocation | Defining a study period at least 3 times the maximum temporal window helps with statistical power. Extending the study period further may decrease the accuracy of the temporal trend adjustment but might be of interest to detect more prolonged clusters. If citywide percent positivity reaches an inflection point (e.g., begins to increase again after a period of decrease), the study period will need to be temporarily shortened and reset after that inflection point to accurately adjust for the temporal trend. For longer temporal window, selected June 1, 2020 as earliest date when citywide percent positivity trend appeared stable without an inflection point. |
| Lag for data accrual | 3 days | Given lags between specimen collection and report, exclude very incomplete data at end of study period when estimating the temporal trend. Three days is the minimum lag possible to preserve a timely analysis while allowing for at least some data to be reported, geocoded, and analyzed prior to open of business. |

\*Turner K, Davidson SL, Collins J, Park SY, Pedati CS. Council of State and Territorial Epidemiologists (CSTE) standardized surveillance case definition and national notification for 2019 novel coronavirus disease (COVID-19) (https://cdn.ymaws.com/www.cste.org/resource/resmgr/2020ps/Interim-20-ID-01_COVID-19.pdf). 2020.
†See Technical Appendix.
‡Levin-Rector A, Nivin B, Yeung A, Fine AD, Greene SK. Building-level analyses to prospectively detect influenza outbreaks in long-term care facilities: New York City, 2013-2014. *Am J Infect Control*. 2015;43(8):839-843.

Given changing cluster definitions and investigation protocols during the study period, instead of summarizing all clusters detected, we present three illustrative examples (Table 2). On June 22, in the context of waning case counts citywide, the system detected a not statistically significant cluster of 6 patients residing in a 0.6-kilometer radius, all with specimens collected on June 17 (Figure 1A). DOHMH staff interviewed patients for common exposures, such as attending the same event or visiting the same location. On June 23, a DOHMH surveillance investigator (D.B.) determined that two patients in the cluster had attended the same gathering, where recommended social distancing practices had not been observed. In response, DOHMH launched an effort to limit further transmission, including testing, contact tracing, community engagement, and health education emphasizing the importance of isolation and quarantine. Investigation of patients in the cluster with the highest recurrence interval (323 days) during the study period did not reveal any epidemiologic linkages. Detection of a sustained cluster (lasting >1 week) with high percent positivity (8.9%) (Figure 1B) supported the selection of one ZIP code for targeted testing and outreach, as part of NYC’s hyper-local plan to prevent COVID-19 transmission (*14*).

**Table 2.**
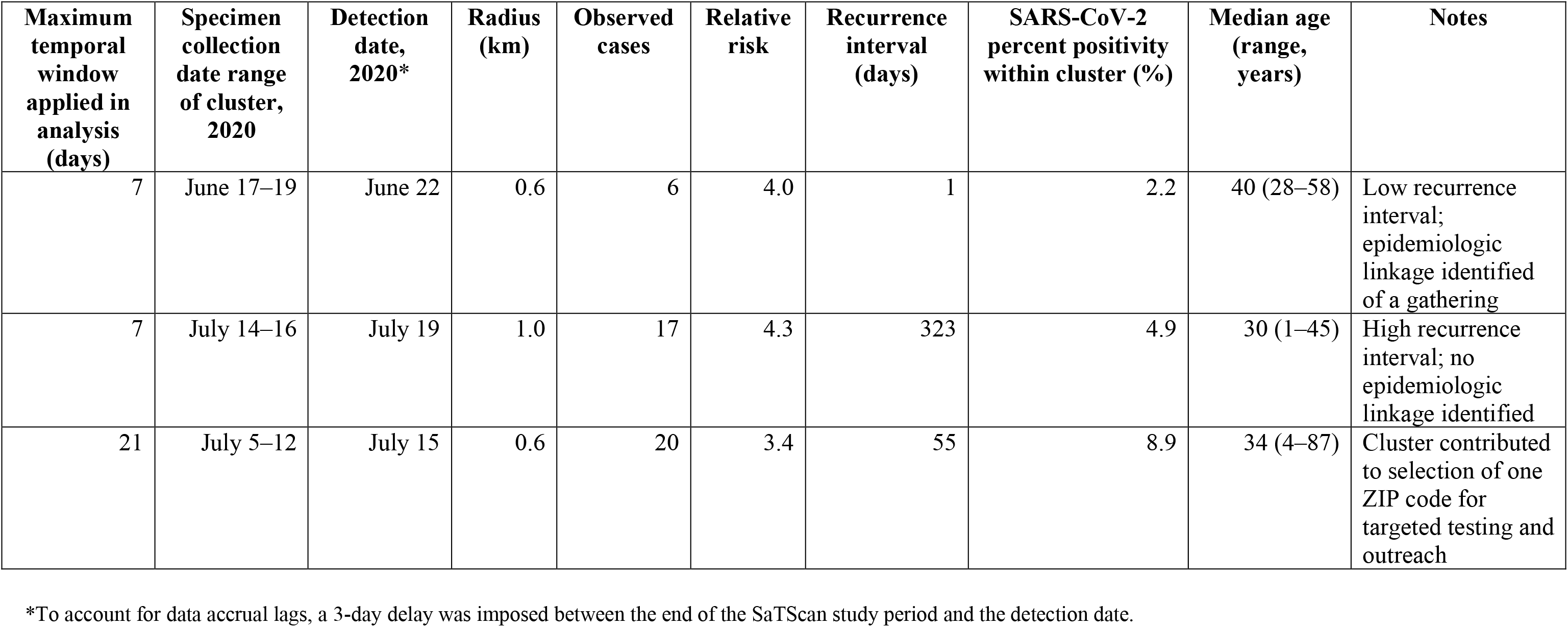
Selected SARS-CoV-2 percent positivity clusters prospectively detected during June–July 2020, New York City.

| Maximum temporal window applied in analysis (days) | Specimen collection date range of cluster, 2020 | Detection date, 2020* | Radius (km) | Observed cases | Relative risk | Recurrence interval (days) | SARS-CoV-2 percent positivity within cluster (%) | Median age (range, years) | Notes |
| --- | --- | --- | --- | --- | --- | --- | --- | --- | --- |
| 7 | June 17–19 | June 22 | 0.6 | 6 | 4.0 | 1 | 2.2 | 40 (28–58) | Low recurrence interval; epidemiologic linkage identified of a gathering |
| 7 | July 14–16 | July 19 | 1.0 | 17 | 4.3 | 323 | 4.9 | 30 (1–45) | High recurrence interval; no epidemiologic linkage identified |
| 21 | July 5–12 | July 15 | 0.6 | 20 | 3.4 | 55 | 8.9 | 34 (4–87) | Cluster contributed to selection of one ZIP code for targeted testing and outreach |

\*To account for data accrual lags, a 3-day delay was imposed between the end of the SaTScan study period and the detection date.

**Figure 1.**
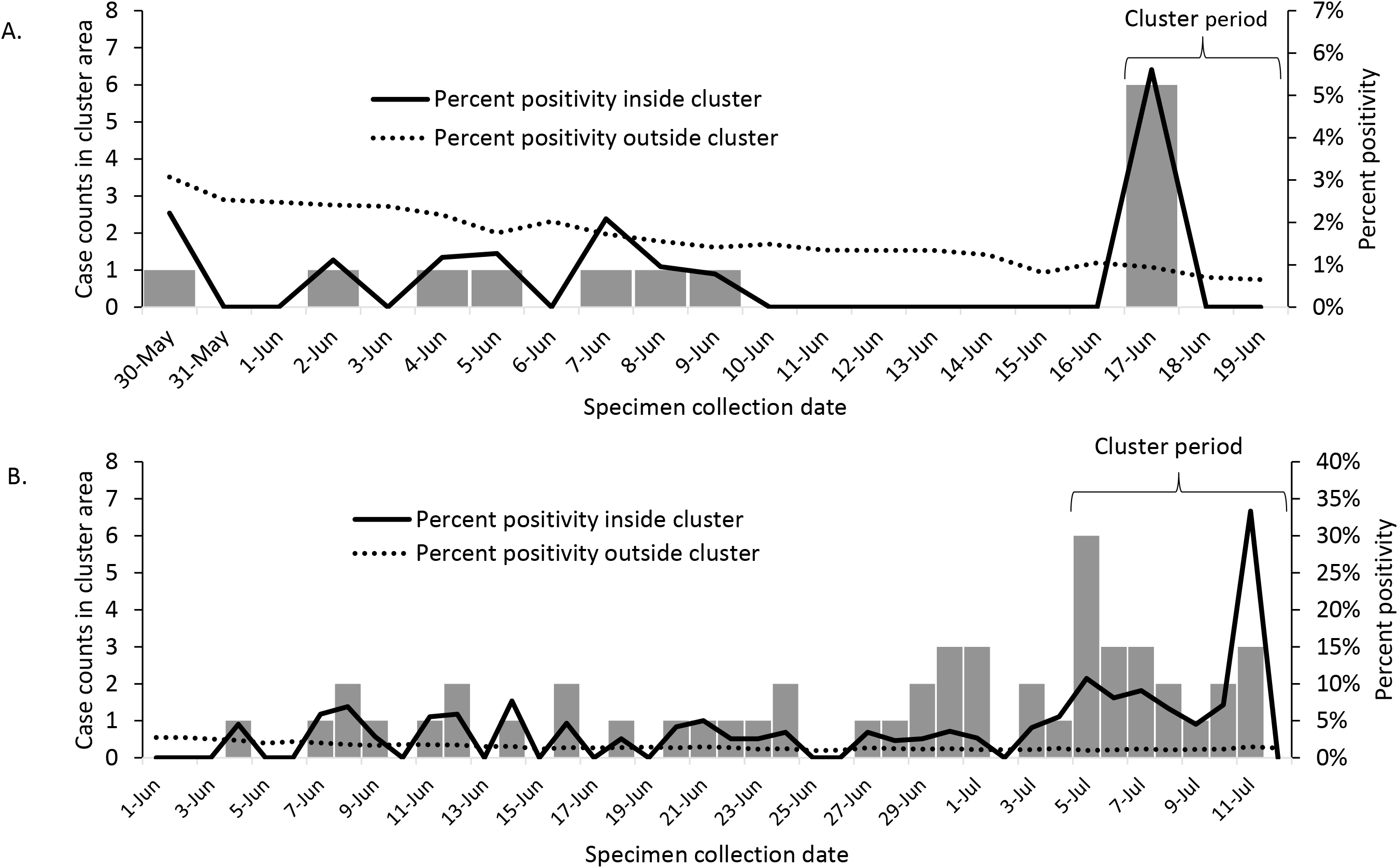
Cluster case counts and SARS-CoV-2 percent positivity inside and outside cluster area for clusters detected in New York City (A) on June 22, 2020 in 5 census tracts, in which patients reported common attendance at a social gathering, and (B) on July 15, 2020, in 7 census tracts, contributing to the selection of one ZIP code for targeted testing and outreach.

## Conclusions

Automated spatiotemporal cluster detection analyses detected emerging, highly focused areas to target COVID-19 containment efforts in NYC. For jurisdictions where case investigation capacity is limited, interviewing patients in clusters, even if not statistically significant, can help identify common exposures for interrupting further transmission. Regardless of whether epidemiologic linkages are identified — or whether overall transmission is increasing, decreasing, or steady — clusters can be used to prioritize resources and focus interventions such as promoting increased testing, public messaging, and community engagement activities.

Our system is subject to several limitations. First, analyses were based on specimen collection date, but given delays in testing availability and care seeking, these dates did not necessarily represent recent infections. Timeliness was further limited by delays from specimen collection to laboratory testing and reporting. Clusters dominated by asymptomatic patients or patients with illness onset >14 days prior to diagnosis may not require intervention, as a positive PCR result indicates the presence of viral RNA but not necessarily viable virus (*15*). Second, geocoding is required for precision, and of unique NYC residents with a specimen collected during June–July 2020 for a PCR test for SARS-CoV-2 RNA, approximately 3% had a non-geocodable residential address and were excluded from analyses. Finally, automation coding was complex (Technical Appendix). Planned SaTScan software enhancements that will facilitate wider adoption by other health departments include: adding a software interface for prospective surveillance, enabling temporal and spatial adjustments for the Bernoulli probability model, and enabling the log-linear temporal trend adjustment with automatically calculated trend at a sub-annual scale.

Our COVID-19 early detection system has highlighted areas in NYC warranting a rapid response. Such local targeted, place-based approaches are necessary to minimize further transmission and to better protect people at high risk for severe illness, including older adults and people with underlying health conditions.

## Data Availability

Data at high spatiotemporal resolution are not publicly available in accordance with patient confidentiality and privacy laws. Publicly available data are linked below.

https://www1.nyc.gov/site/doh/covid/covid-19-data.page

## Acknowledgments

We thank all staff members of the DOHMH Incident Command System Surveillance and Epidemiology Section for processing, cleaning, and managing input data; for conducting patient interviews and cluster investigations; and for logistical support. We also thank the NYC Test and Trace Corps for their assistance in managing the cases and contacts included in and identified by cluster investigations.S.K.G. and E.R.P were supported by the Public Health Emergency Preparedness Cooperative Agreement (grant NU90TP922035-01), funded by the Centers for Disease Control and Prevention. This article’s contents are solely the responsibility of the authors and do not necessarily represent the official views of the Centers for Disease Control and Prevention or the Department of Health and Human Services.

## First author biographical sketch

Dr. Greene is the director of the Data Analysis Unit at the Bureau of Communicable Disease of the New York City Department of Health and Mental Hygiene, Long Island City, New York. Her research interests include infectious disease epidemiology and applied surveillance methods for outbreak detection.

## Technical Appendix

**Technical Appendix Table 1.** Analysis parameter settings for SARS-CoV-2 percent positivity analyses in New York City, using the prospective Poisson-based space-time scan statistic.

| Parameter | Parameter setting | Notes |
| --- | --- | --- |
| Analysis type | Prospective space-time | For timely cluster detection, prospective (rather than retrospective) analyses are used, evaluating only the subset of possible clusters that encompass the last day of the study period. To detect acute, ongoing, localized disease clusters, space-time analyses (rather than purely temporal or purely spatial analyses), are used |
| Model type | Discrete Poisson | We apply the discrete Poisson-based scan statistic, defining the “population” file as persons tested, to scan for clusters of increased percent positivity. If SARS-CoV-2 percent positivity is high (say, >10%), then the discrete Poisson-based scan statistic is a poor approximation for Bernoulli-type data of persons testing positive and negative. The analysis would produce conservative p-values (i.e., recurrence intervals biased too low), and true clusters might be missed. However, SaTScan v9.6 does not include features for spatial and temporal adjustments for the Bernoulli probability model. |
| Maximum spatial cluster size | 50% of the population being tested | The option that imposes the fewest assumptions is to allow the cluster to expand in size to include up to 50% of all persons tested during the study period. Forcing clusters to be smaller than 50%, or restricting in terms of geographic size by setting a maximum circle radius, can be motivated in geographically larger study regions. |
| Maximum temporal cluster size | 7 days* for analysis to support prioritization of case investigations; 21 days for analysis to support place-based resource allocation | To focus on hotspots emerging during the most recent week; to focus on areas with more sustained emerging increases. |
| Minimum temporal cluster size | 3 days* for analysis to support prioritization of case investigations; 7 days* for analysis to | Clusters of <3-day duration considered less credible for investigation as an emerging hotspot; clusters of <7 days considered lower priority for resource allocation. |
|  | support place-based resource allocation |  |
| Minimum number of cases | 5 cases | Require a minimum number of cases to improve the probability of at least 3 patients within a given cluster being reachable for interview to support identification of a common exposure. |
| Temporal trend adjustment | Log-linear with automatically calculated trend* | If citywide percent positivity decreasing overall, then wish to detect areas where decreasing slower than citywide average. If citywide percent positivity increasing overall, then wish to detect areas where increasing more than citywide average. Adjusting for temporal trend nonparametrically is not possible if also using nonparametric spatial adjustment. |
| Spatial adjustment | Nonparametric, with spatial stratified randomization | Goal is to detect areas with relative increases from baseline, even if still lower than average citywide. This method adjusts the expected count separately for each location, removing all purely spatial clusters. The randomization is then stratified by location ID to ensure that each location has the same number of events in the real and random data sets. |
| Scan for areas with: | High rates | Interested only in increased disease transmission. |
| Inference | Default p-value method, with maximum number of Monte Carlo replications = 9999 | A maximum of 9999 replications increases power compared with 999 replications and is computationally feasible. |
| Secondary cluster reporting criteria (output parameter) | No cluster centers in other clusters | Any disease may have multiple active clusters at any moment, so secondary clusters should be reviewed. By reviewing clusters with no cluster centers in other clusters (rather than no, or more geographic overlap), secondary clusters with some overlap can be detected. |

\* See “study period and time precision” section below.

### Geocoding

Patient addresses were geocoded daily using version 20A of the NYC Department of City Planning’s Geosupport geocoding software, implemented in R through C++ using the Rcpp package.^3^ Addresses that failed to geocode were then cleaned using a string searching algorithm performed against the Department of City Planning’s Street Name Dictionary and Property Address Directory. Addresses that failed to geocode after cleaning were then verified using the IBM Infosphere USPS service.

### Study period and time precision

SaTScan v.9.6 can estimate a temporal trend (see below), but only at an annual time scale, as this feature was originally developed to accommodate long-term secular trends across multiple years, as for cancer incidence. As a workaround to accommodate a rapidly changing trend, as for SARS-CoV-2 test positivity, reassign one day as if it were one year in the SaTScan case and population input files and conduct analyses at annual resolution. For example, for a 21-day study period ending June 19, 2020, reassign May 30, 2020 as the year “2000” and June 19, 2020 as the year “2020,” and indicate a time precision and a time aggregation of “year,” (i.e., PrecisionCaseTimes=1 and TimeAggregationUnits=1 in the SaTScan parameter file). The minimum and maximum temporal cluster sizes would be input as years instead of days. Similarly, with input data expressed in years, nonparametric adjustment for space by day-of-week interaction was not possible. Note that in calculating the recurrence interval, SaTScan assumes that analyses are repeated on a regular basis with a periodicity equal to the specified time interval length. Because these daily analyses are specified at annual time intervals, interpret recurrence intervals in days, not years; e.g., a recurrence interval of 1.0 years in this context should be re-interpreted as 1.0 days, i.e., consistent with chance alone.

### Temporal trend adjustment

As a workaround for a bug in SaTScan v.9.6 in calculating a temporal trend adjustment in the prospective setting, first use the case and population files to run a retrospective purely temporal Poisson analysis, with the temporal adjustment “Log linear with automatically calculated trend” (TimeTrendAdjustmentType=3 in the SaTScan parameter file). Read in this automatically calculated temporal trend from the SaTScan text output. Retain the magnitude of trend (“X”) and sign of X determined by “increase” or “decrease.” Example SaTScan text output excerpt:

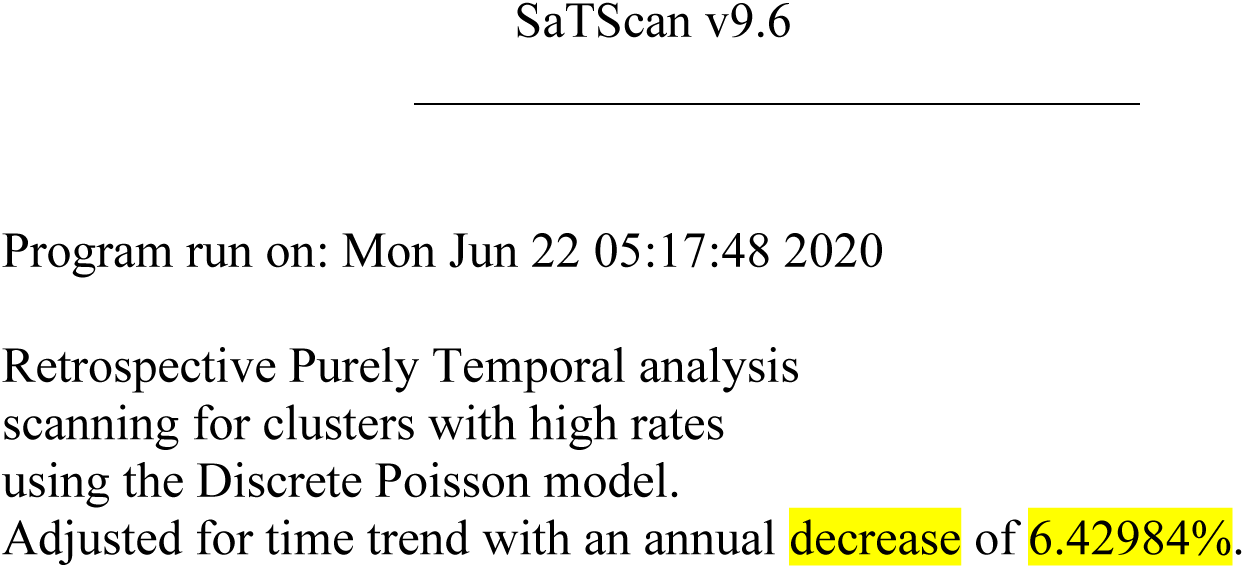

The time trend is the same for retrospective and prospective analyses. Then, run the prospective spatio-temporal Poisson analysis, inserting the calculated time trend in the parameter file as user-specified (TimeTrendAdjustmentType=2, TimeTrendPercentage=-6.42984 in the SaTScan parameter file). Example user interface screenshot:

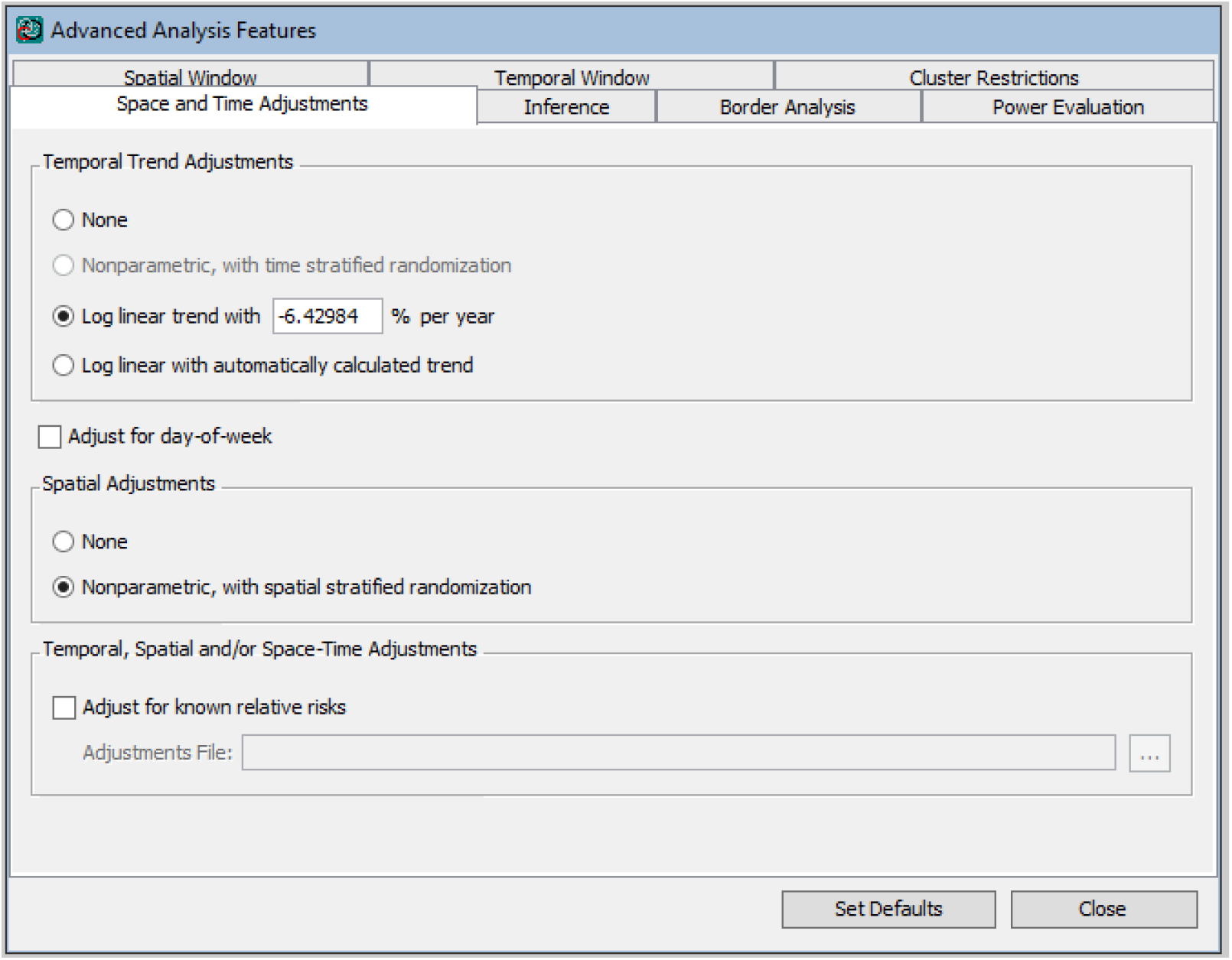

If unable to enter a negative value for the temporal trend adjustment in the user interface, then edit the “TimeTrendPercentage” value in the SaTScan parameter file in a text editor (e.g., Notepad), then reopen in SaTScan.

## Footnotes

1 Kulldorff M, Information Management Services, Inc. SaTScan v9.6: software for the spatial and space-time scan statistics (www.satscan.org). 2018.

2 https://github.com/CityOfNewYork/communicable-disease-surveillance-nycdohmh

3 Eddelbuettel D, Francois R. Rcpp: Seamless R and C++ integration. *Journal of Statistical Software*. 2011;40(8):1–18.

